# Image Quality Evaluation of Neonatal Brain MRI Using a Deep Learning Reconstruction Algorithm: A Quantitative and Multireader Study Using Variable Denoising Levels at 3 Tesla

**DOI:** 10.64898/2026.02.04.26345479

**Authors:** Zain Alvi, Eduardo P. Reis, David D. Shin, Suchandrima Banerjee, Hisham M. Dahmoush, Andrew Campion, Mateus A. Esmeraldo, Stefanie Chambers, Yanniklas Kravutske, Sergios Gatidis, Bruno P. Soares

## Abstract

**Purpose:** Neonatal imaging is particularly challenging because newborns have a high likelihood of head motion, which can degrade image quality and complicate interpretation. Improving MRI brain image quality may help reduce diagnostic uncertainty and facilitate the nuanced assessment of early myelinating structures in the neonatal brain. Although deep learning reconstruction algorithms designed to improve MRI image quality have been evaluated in pediatric imaging, they have not been specifically studied in exclusively neonatal populations. We sought to evaluate image quality improvement through the employment of a deep learning reconstruction algorithm in neonatal brain imaging.

**Methods:** 3D T1-weighted brain MRIs were obtained in 15 neonates. A deep-learning reconstruction algorithm was applied to the image sets using low, medium, and high levels of denoising. Three radiologists qualitatively rated image quality (signal-to-noise ratio, presence of artifacts, and overall clarity) on a 4-point scale of eight early myelinating structures. Objective apparent signal-to-noise ratio (aSNR) and apparent contrast-to-noise ratio (aCNR), based on signal intensities of white-and gray-matter, was measured across all three denoising levels.

**Results:** Evaluation by radiologists indicated an overall increase in all image quality categories and increased conspicuity of the early myelinating structures as the level of denoising increased. Objective aSNR and aCNR values also increased progressively with denoising, with significant differences observed for nearly all pairwise comparisons.

**Conclusion:** Our findings suggest that the use of the proposed deep learning reconstruction algorithm improves image quality in 3D T1-weighted neonatal brain MRIs at 3T.

## Introduction

Neonatal brain MR imaging is challenging due to the high probability of head motion since it is usually performed with little or no sedation [1]. Motion during image acquisition causes artifacts that may affect the diagnostic capability and confidence of the interpreting radiologist [1,2]. Therefore, fast and motion-robust imaging techniques are desirable, as they may help reduce uncertainty when image quality is compromised [3]. Multiple strategies have been developed to shorten MRI acquisition times, including parallel imaging compressed sensing, and radial k-space sampling. While these techniques can substantially accelerate imaging, they often come at the cost of reduced signal-to-noise ratio (SNR), residual aliasing or reconstruction artifacts, and occasionally blurring or altered image texture that can obscure subtle pathology [4].

In neonatal brain MR imaging, employing a 3D Gradient Echo (3D GRE) T1-weighted sequence is preferable to depict early myelinating brain structures, as myelin appears hyperintense on T1-weighted images [5]. The 3D acquisition also provides higher anatomical details, and the flexibility to view the structures in any spatial orientation via reformats. Early myelinating structures are key brain areas to assess as the presence of myelin correlates with functionality. Recently, deep learning reconstruction techniques have shown promise in image quality enhancement by reducing noise, improving signal-to-noise ratio, and allowing for shorter image acquisition times [6]. Traditional motion-compensating techniques are available and can be used in tandem with deep-learning reconstruction to provide higher-quality images and make neonatal neurological diagnoses more reliable [7,8].

A specific denoising tool, AIR Recon DL, has shown promise in improving image quality [9]. The AI model was trained to minimize random image noise and Gibbs ringing artifact (also known as truncation artifact) in complex MRI data based on ideal images. By substantially improving signal-to-noise ratio and edge sharpness while reducing Gibbs ringing artifacts, the algorithm can enable shorter scan times by allowing operators to optimize the acquisition protocol. Shorter acquisition times can be achieved by reducing the number of signal averages, increasing acquisition bandwidth, or applying higher parallel imaging acceleration. Although the algorithm does not directly correct motion-related artifacts, they may be indirectly reduced by those acquisition protocol optimizations that lead to shorter scan times [9].

AIR Recon DL is a raw-data–based MR reconstruction integrated into the scanner pipeline; it uses a convolutional neural network (4.4 million parameters across around 10,000 kernels) with rectified linear unit activation activations and no bias terms to perform blind denoising [9]. It was trained on more than 10,000 near-perfect and conventional datasets spanning varied contrasts and shapes and using diverse augmentations (e.g. rotation, Gaussian noise, phase manipulation) [10]. The algorithm was designed to function with any patient and pathology [9].

While this tool has been used to optimize adult neurological MRI exams in cases of tumor identification, and although it has been tested in the pediatric population, it has not yet been specifically investigated in neonatal brain MRI [6, 11]. This gap is notable given the unique technical and diagnostic demands of neonatal neuroimaging. Accordingly, the goal of this investigation is to assess the ability of AIR Recon DL to improve image quality in 3D T1-weighted neonatal brain imaging at 3T.

## Materials and Methods

### Subjects

The study was designed as a retrospective case series. A consecutive sample of 15 neonatal scans acquired in May 2024 was selected, irrespective of image quality. Preliminary analyses demonstrated highly consistent and statistically significant results across all evaluated categories, indicating stable and reproducible patterns in the data, as detailed in the results section. Given this level of consistency, the inclusion of additional patients would not be expected to yield new insights or alter the observed trends. Therefore, expanding the dataset further was considered unnecessary for the objectives of the present study.

These patients underwent clinically indicated T1-weighted MRI at a single medical center. These neonates, who required MRI as part of their clinical care, had no specific exclusion criteria. The institutional review board approved the protocol. The imaging findings of each patient, along with basic demographic data, are summarized in **Table 1**.

**Table 1.** Summary demographics and imaging characteristics of the neonatal cohort.

| **Variable** | **Count** |
| --- | --- |
| **Number of neonates** | 15 |
| **Sex** | 11 male, 4 female |
| **Postnatal age at MRI** | 0–7 days (n = 9); 8–14 days (n = 4); 15–28 days (n = 2) |
| **Imaging findings** | Normal (n = 2); White matter signal abnormalities (n = 5); Hemorrhagic findings (n = 4); Ventriculomegaly or structural abnormalities (n = 4) |
| **MRI indication** | Suspected hypoxic–ischemic or ischemic injury (n = 4); suspected intracranial hemorrhage (n = 5); ventriculomegaly or CSF space abnormalities (n = 3); suspected congenital/developmental anomalies (n = 2); screening with no significant abnormality (n = 1) |

### Procedure

All MRI examinations were performed on 3T GE HealthCare scanners (SIGNA Premier, Discovery MR750, SIGNA PET/MR, and SIGNA Architect; GE HealthCare, Waukesha, WI) using standard head coils. The acquisition parameters for the T1-weighted axial sequence were: TR = 7.3 ms, TE = 3 ms, TI = 600 ms, flip angle = 8°, NEX = 1, spatial resolution = 0.9 × 0.9 × 1 mm^3^, bandwidth = 244 Hz/pixel, acceleration = 1.5×1, and scan time = 4:30 min.

Subsequently, three additional image sets were generated in DICOM format for each exam using the AIR Recon DL deep learning reconstruction algorithm from the raw k-space data for each patient at low, medium, and high denoising levels [9].

In the qualitative analysis, three board-certified radiologists (one pediatric neuroradiologist with 20 years of experience, one pediatric radiologist with 10 years of experience, and one neuroradiology fellow) were recruited to assess each image. The image sets were anonymized and presented in a random arrangement, blinded to the readers. The sets included the original acquisition and images processed with low, medium, and high levels of denoising.

Radiologists evaluated the images on a 1 to 4 scale (1 = worst, 4 = best) for perceived signal-to-noise ratio (SNR), overall image quality (OIQ), and the presence of additional artifacts (ADA) such as artifacts related to motion, field inhomogeneity, and ghosting. In the ADA assessment, radiologists focused on evaluating artifacts other than those primarily addressed by the algorithm (Gibbs ringing and random image noise). These two artifacts are the main targets of the deep-learning reconstruction software, based on how it was trained and designed, as described in the introduction section [9]. Their reduction is expected to be partially reflected in both the SNR and OIQ scores.

Data collection also involved radiologists rating the conspicuity and confidence in evaluating early myelinating structures, including the medulla, dorsal pons, decussation of superior cerebellar peduncles, inferior colliculi, subthalamic nuclei, globi pallidi, posterior limb of the internal capsules, and periolandic cortices, using the same 1-4 ranking scale.

For quantitative analysis, two circular 5 mm regions of interest (ROIs) were placed in the right frontal white matter (WM) and right putamen using 3D Slicer, following methodology previously validated, without modification [12, 13, 14]. The placement was performed by consensus between two board-certified radiologists (an expert pediatric neuroradiologist and a postdoctoral research fellow in pediatric neuroradiology). Quantitative metrics were extracted within both ROIs on a per case and denoising level-basis, with which following measures were calculated: apparent signal-to-noise ratio (aSNR) and apparent contrast-to-noise ratio (aCNR), based on signal intensities of white-and gray-matter [13].

### Terms and Measures

Perceived Signal-to-Noise Ratio: The radiologist’s perception of the clarity of the image in differentiating signal from noise.

Presence of Additional Artifacts: Any additional artifacts present in the image other than Gibbs ringing that could limit diagnostic ability or confidence, such as motion artifacts and field inhomogeneity.

Overall Image Quality: Radiologist’s overall evaluation of the image based on a combination of perceived SNR, sharpness, and the absence of artifacts.

Apparent Signal-to-Noise Ratio (aSNR): Calculated based on the local variability within the tissue region of interest (ROI).

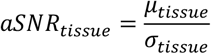

where μ is the mean signal intensity and σ is the standard deviation within the ROI [13]. Apparent Contrast-to-Noise Ratio (aCNR): Calculated as the ratio of the difference in mean signal intensities between white matter (WM) and gray matter (GM) to the combined standard deviations in their respective ROIs.

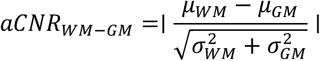

where μ is the mean signal intensity and σ is the standard deviation for the respective tissue type [13].

### Statistical Tests

Mean reader scores were evaluated for each image. Individual segment scores for early myelinating structures were separately calculated. A non-parametric repeated-measures analysis was conducted using the Aligned Rank Transform (ART) ANOVA to assess the effect of DL reconstruction strength on rating scores, while controlling for Patient, Reader, and Category as random effects. The model was fit using the ARTool package in R, and Type III Wald F-tests were used to evaluate main effects. Post hoc pairwise comparisons between reconstruction levels were performed using Tukey-adjusted contrasts on the aligned ranks via the emmeans package. P-values < 0.05 were considered statistically significant.

aSNR in GM and WM, as well as aCNR (WM–GM), were evaluated across subjects at each denoising level (Original, Low, Medium, High). Subject-level data were aggregated, and per-condition means, standard deviations, sample sizes (N), and standard errors (SE = SD/√N) were computed. Values were visualized as mean ± SE. To formally assess whether denoising level significantly influenced quantitative metrics, a one-way repeated-measures ANOVA was conducted for each group (aSNR in GM, aSNR in WM, and aCNR), with denoising level as the within-subject factor (N=15). Where the omnibus ANOVA was significant, post-hoc pairwise comparisons were performed using paired t-tests with Holm correction for multiple comparisons. Results were presented as grouped bar plots with SE error bars. All analyses and figures were produced in Python (version 3.13.15) using pandas, numpy, and matplotlib, and custom scripts with SciPy/Statsmodels for statistical testing.

## Results

The scores for the 15 neonatal brain MRIs for each patient with various denoising levels are presented below. **Table 2** shows the scores for signal-to-noise ratio, presence of additional artifacts, and overall image quality. **Table 3** shows the scores of the individual rankings for the visibility of the different early myelinating structures.

**Table 2.** Mean image quality reader evaluation scores and standard deviation for signal-to-noise ratio (SNR), presence of additional artifacts (ADA), and overall image quality (OIQ) for all image sets at the low, medium, and high denoising settings compared to the original acquisition.

| Category | Original | Low | Medium | High |
| --- | --- | --- | --- | --- |
| SNR | 2.20 ± 0.68 | 2.67 ± 0.62 | 3.13 ± 0.52 | 3.60 ± 0.51 |
| ADA | 2.80 ± 0.86 | 2.93 ± 0.88 | 3.20 ± 0.77 | 3.53 ± 0.74 |
| OIQ | 2.20 ± 0.68 | 2.53 ± 0.64 | 2.93 ± 0.59 | 3.53 ± 0.52 |

**Table 3.** Mean image quality score of early myelinating structures [globi pallidi (GP), inferior colliculi (IC), medulla (MED), perirolandic cortices (PC), posterior limb of the internal capsules (PLIC), dorsal pons (PONS), decussation of superior cerebellar peduncles (SCP), subthalamic nuclei (SN)], at low, medium, and high levels of denoising compared to the original acquisition with standard.

| Category | Original | Low | Medium | High |
| --- | --- | --- | --- | --- |
| GP | 2.80 ± 0.56 | 2.80 ± 0.56 | 3.20 ± 0.56 | 3.87 ± 0.35 |
| IC | 2.73 ± 0.59 | 2.93 ± 0.80 | 3.40 ± 0.91 | 3.67 ± 0.82 |
| MED | 2.73 ± 0.59 | 2.73 ± 0.70 | 3.33 ± 0.90 | 3.67 ± 0.62 |
| PC | 2.47 ± 0.52 | 2.67 ± 0.72 | 2.93 ± 0.59 | 3.47 ± 0.52 |
| PLIC | 2.53 ± 0.74 | 2.67 ± 0.72 | 3.07 ± 0.80 | 3.73 ± 0.80 |
| PONS | 2.73 ± 0.59 | 3.00 ± 0.65 | 3.13 ± 0.52 | 3.73 ± 0.59 |
| SCP | 2.67 ± 0.72 | 2.93 ± 0.70 | 3.33 ± 0.82 | 3.67 ± 0.82 |
| SN | 2.80 ± 0.77 | 2.87 ± 0.83 | 3.27 ± 0.70 | 3.80 ± 0.56 |

The ART ANOVA revealed a significant main effect of DL reconstruction strength on reader scores (p < 0.0001), indicating that DL reconstruction strength significantly influenced ratings even after accounting for variability due to individual patient, reader, and category. Post hoc comparisons showed that all reconstruction levels differed significantly from each other (Tukey-adjusted p < 0.0001), with scores increasing consistently from Original < Low < Medium < High. Specifically, there was a strong positive association between increased denoising levels and increased ratings in signal-to-noise ratio, presence of additional artifacts, and overall image quality (**Figure 1**). A similar association was found between visibility of early myelinating structures and denoising strength settings (**Figure 2**).

**Figure 1:**
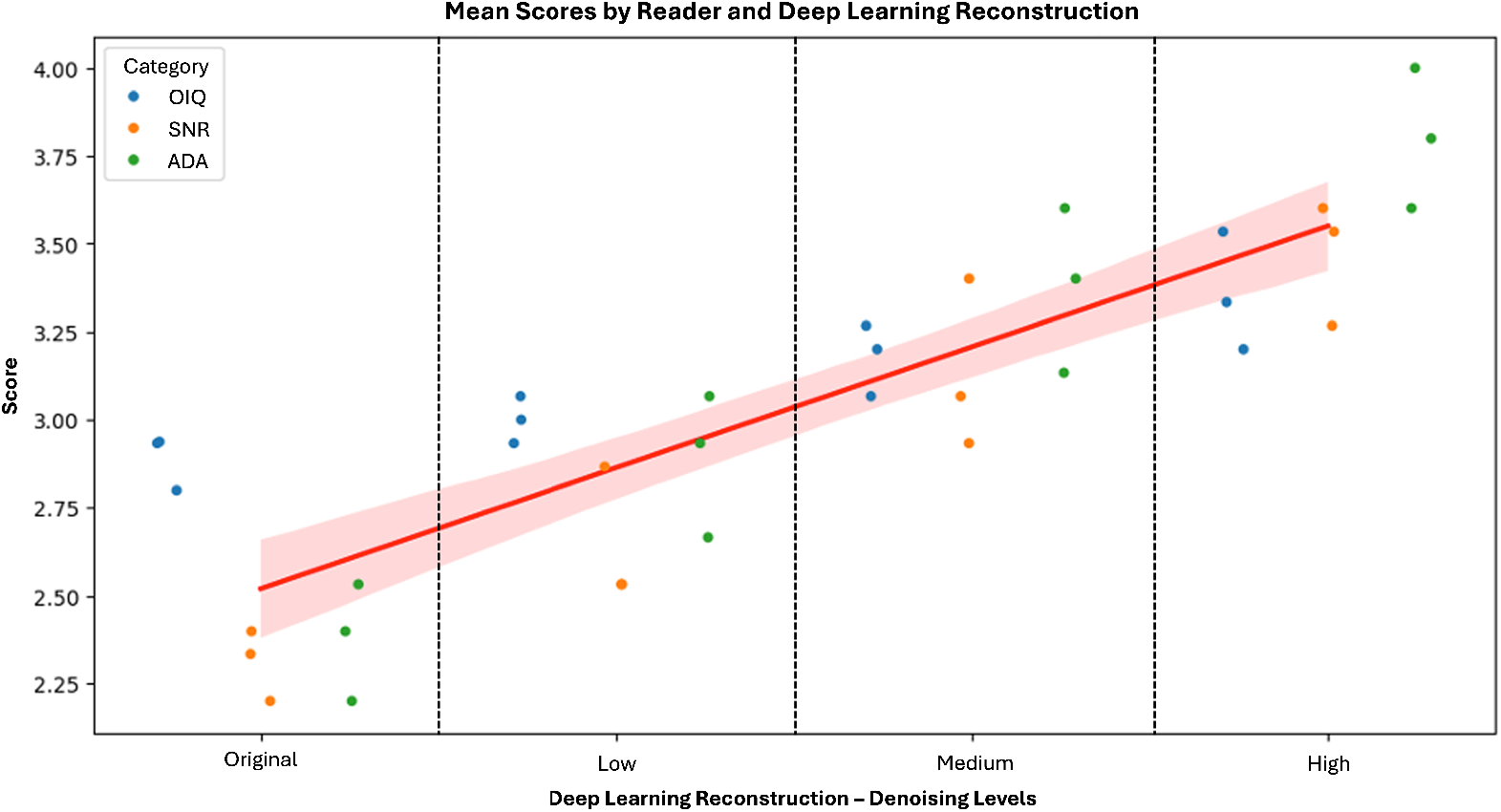
Mean overall quality scores [overall image quality (OIQ), signal-to-noise ratio (SNR), and presence of additional artifacts (ADA)] by reader at low, medium, and high denoising levels compared to the original acquisition.

**Figure 2:**
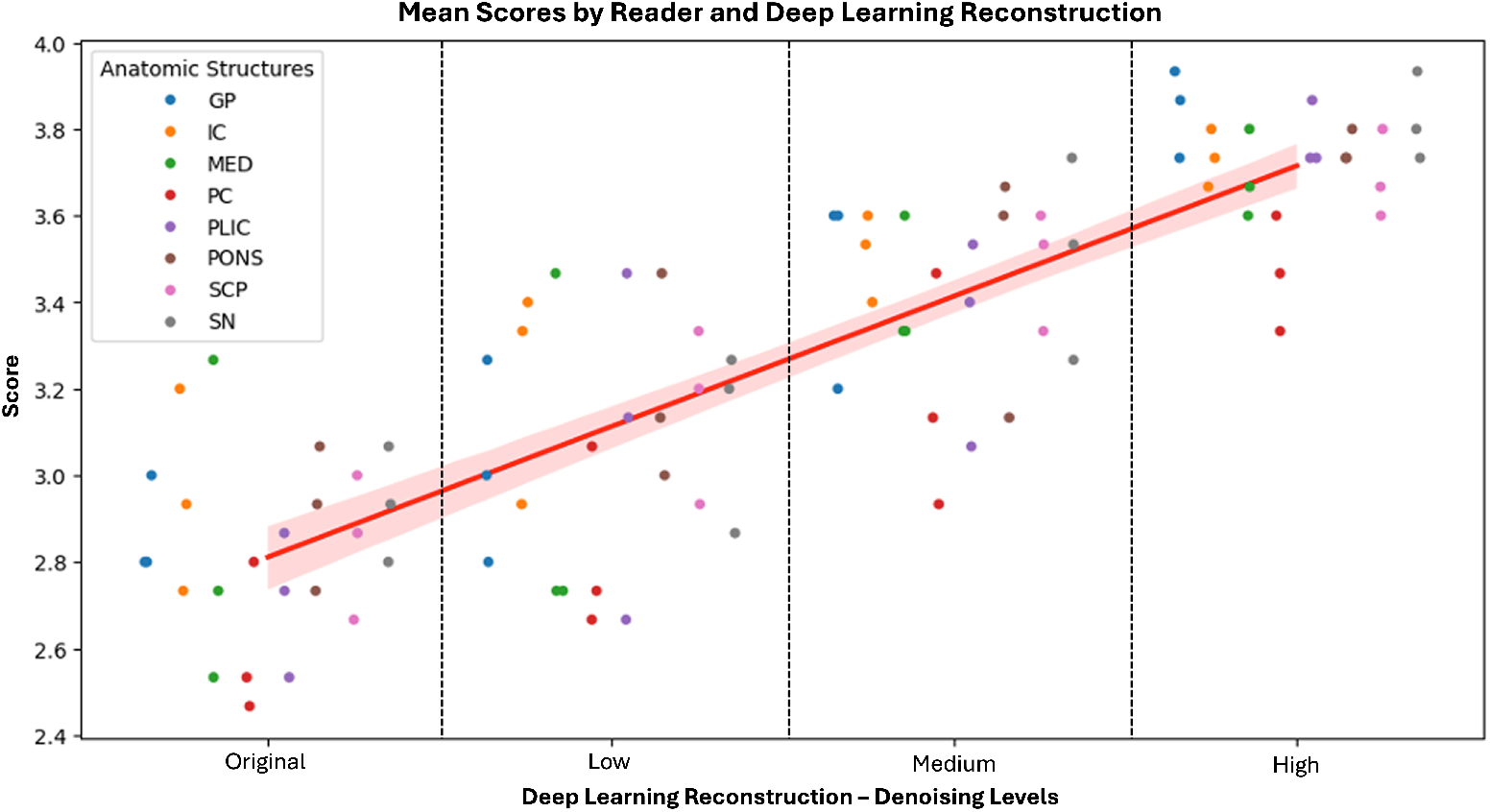
Mean overall quality scores of early myelinating structures [globi pallidi (GP), inferior colliculi (IC), medulla (MED), perirolandic cortices (PC), posterior limb of the internal capsules (PLIC), dorsal pons (PONS), decussation of superior cerebellar peduncles (SCP), and subthalamic nuclei (SN)] at low, medium, and high levels of denoising compared to the original acquisition.

Quantitative analysis further supported the qualitative reader evaluations, demonstrating significant improvements in aSNR and aCNR with increasing denoising levels (Table 4 and Figure 6). All values are reported as mean ± standard error (SE). Mean gray matter aSNR increased stepwise from 11.77 ± 1.07 in the original images to 21.73 ± 1.82 at the high denoising setting. Similarly, white matter aSNR rose from 6.48 ± 0.71 (original) to 12.91 ± 1.07 (high). Apparent CNR between WM and GM also improved consistently, increasing from 3.96 ± 0.31 at baseline to 7.38 ± 0.55 with high denoising. All pairwise comparisons were statistically significant (p ≤ 0.04), with the exception of WM aSNR between the original and the low level of denoising.

**Table 4.** Quantitative evaluation showed progressive increases in both apparent signal-to-noise ratio (aSNR) and apparent contrast-to-noise ratio (aCNR) with higher denoising levels of the deep learning reconstruction algorithm. Values are reported as mean ± standard error (SE). Mean aSNR in gray matter (GM), white matter (WM), and WM–GM aCNR increased across denoising levels, with all pairwise comparisons statistically significant (p ≤ 0.04) except for WM aSNR between the original and low denoising settings. High-level denoising consistently produced the greatest improvements.

| Metric | Original<br>(Mean ± SE) | Low<br>(Mean ± SE) | Medium<br>(Mean ± SE) | High<br>(Mean ± SE) | Post-hoc comparisons<br>(Holm-corrected) |
| --- | --- | --- | --- | --- | --- |
| aSNR<br>(GM) | 11.77 ± 1.07 | 13.77 ± 1.25 | 16.93 ± 1.48 | 21.73 ± 1.82 | All pairwise differences<br>significant<br>($p < 0.005$) |
| aSNR<br>(WM) | 6.48 ± 0.71 | 7.20 ± 0.51 | 9.23 ± 0.66 | 12.91 ± 1.07 | All significant except<br>Original vs Low |
| aCNR<br>(WM–GM) | 3.96 ± 0.31 | 4.57 ± 0.33 | 5.51 ± 0.48 | 7.38 ± 0.55 | All pairwise differences<br>significant ($p \leq 0.04$) |

## Discussion

The results showed that images that undergo higher denoising levels by the deep learning reconstruction algorithm typically displayed better scores in terms of signal-to-noise ratio, presence of additional artifacts, and overall image quality (**Figures 3-5**), with these qualitative impressions further confirmed by quantitative analysis showing significant improvements in both aSNR and aCNR across all denoising levels (**Figure 6**). The denoising level high, specifically, was the most consistent in receiving the highest scores across all categories. Similarly, the highest level of denoising displayed more clarity in identifying the early myelinating structures, indicating that the AIR Recon DL deep learning reconstruction algorithm improves image assessment of T1-weighted neonatal brain MRIs.

**Figure 3:**
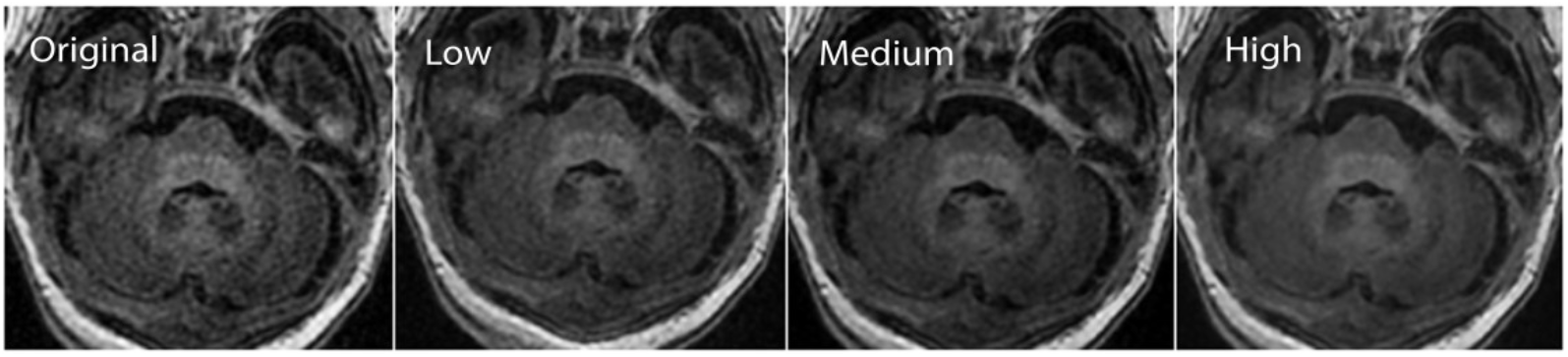
Different levels of deep learning (DL) reconstruction at the level of the pons. Original, low, medium, and high DL denoising algorithms.

**Figure 4:**
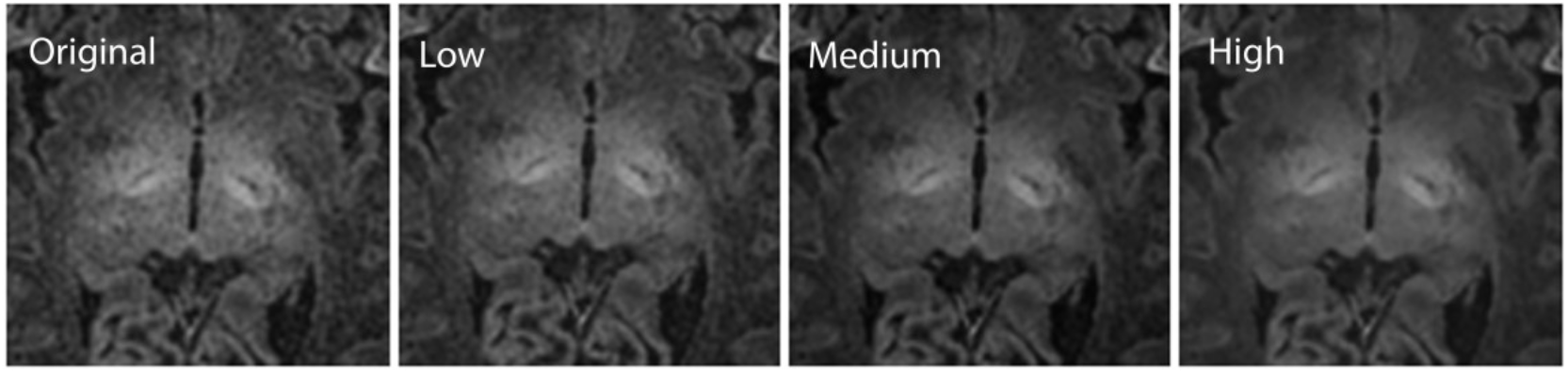
Different levels of deep learning (DL) reconstruction at the level of the subthalamic nuclei and globi pallid. Original, low, medium, and high DL denoising algorithms.

**Figure 5:**
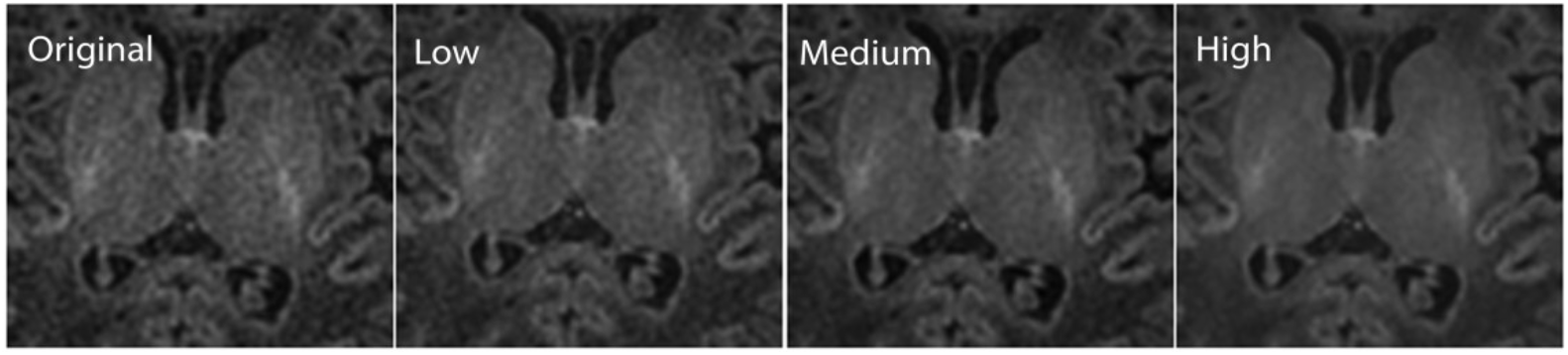
Different levels of deep learning (DL) reconstruction at the level of the posterior limbs of the internal capsules. Original, low, medium, and high DL denoising algorithms.

**Figure 6.**
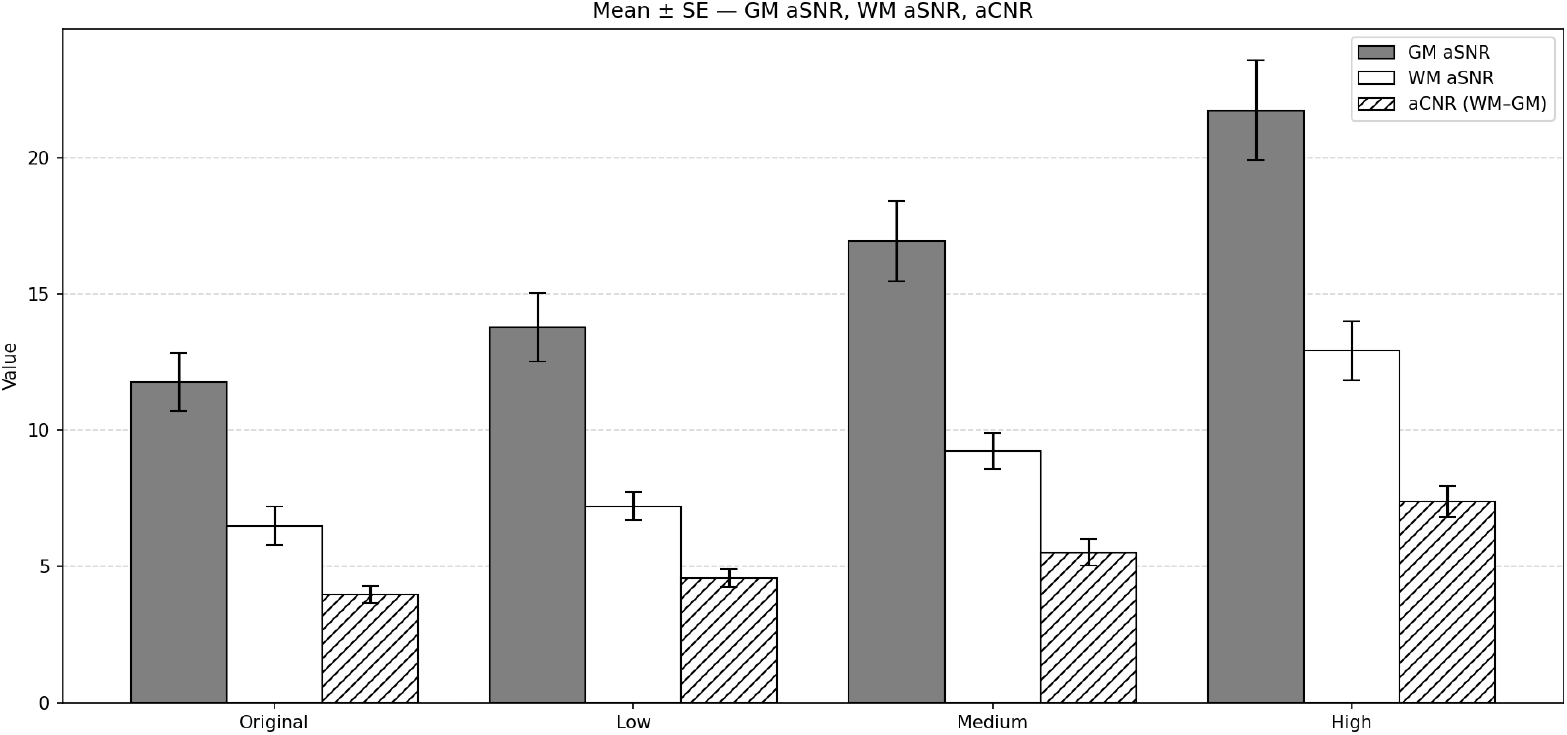
Bar graph showing the mean ± standard error (SE) of apparent signal-to-noise ratio (aSNR) in gray matter (GM, gray bars), aSNR in white matter (WM, white bars), and apparent contrast-to-noise ratio (aCNR, hatched bars) across four denoising levels (Original, Low, Medium, High).

The study demonstrates that the deep learning reconstruction algorithm AIR Recon DL enhances signal-to-noise ratio, overall image quality, and reduces artifacts, facilitating the visualization of early myelinating structures on T1-weighted neonatal brain MRI. Various studies have explored strategies to mitigate MRI motion artifacts. Retrospective motion correction methods have been used in pediatric and adult populations [15,16], but they remain limited in addressing severe or unpredictable neonatal motion [17,18]. Prospective motion correction, while promising, faces technical challenges and high costs [19,20]. In this context, AIR Recon DL enables protocol optimizations that can result in shorter examinations, thereby indirectly reducing the likelihood of motion occurring and offering a valuable complementary solution to these existing challenges. Integrating deep learning reconstruction into motion correction pipelines offers new opportunities to enhance image quality. Deep learning reconstruction algorithms leverage large datasets and complex models to potentially improve diagnostic utility [21,22]. Ongoing development of these tools has the potential to reduce sedation and repeat scans in neonates, improving patient safety and lowering imaging costs [23,24]. In neonatal brain MRI, techniques must balance diagnostic accuracy with practicality and safety. Prior studies stress the importance of minimizing scan duration and motion artifacts, which can compromise diagnostic interpretation [25].

Our findings align with prior research supporting deep learning reconstruction as a tool to enhance image quality [5]. However, several important limitations must be acknowledged. First, because this study focused exclusively on imaging, no clinical outcomes were assessed; consequently, the potential impact of the algorithm on diagnostic decision-making or patient management remains uncertain. Second, although the algorithm may enable protocol optimizations that reduce scan time, these potential reductions were not quantified or assessed in this study. However, such reductions were recently evaluated in pediatric brain MRI using 3D T1 sequences in patients ranging from 5 months to 16 years old, demonstrating acquisition time decreases of 29.3% for pre-contrast and 40.7% for post-contrast scans [14]. Third, we did not evaluate how the algorithm affects the detection of specific diseases, limiting conclusions about improvements in diagnostic performance. Finally, the sample size was small, and although the statistical results were consistent and robust, larger studies will be essential to identify potential unrecognized issues particular to the neonatal population. These factors underscore the need for larger, more diverse studies evaluating algorithm performance in real-world clinical settings.

## Conclusion

In summary, our results show that AIR Recon DL, a deep learning reconstruction technique, improves image quality, signal-to-noise ratio, and visualization of early myelination in neonatal brain MRI at 3T. Future research should focus on quantifying how these improvements translate into real diagnostic and clinical benefits, including whether enhanced image quality can improve diagnostic accuracy for certain diseases, reduce the need for repeat scans due to low image quality, or lessen the need for sedation in neonates.

This study was performed under institutional IRB approval. Ethics and Consent to Participate declarations: All procedures performed in studies involving human participants were in accordance with the ethical standards of the institutional research committee and with the 1964 Helsinki declaration and its later amendments.

## Data Availability

The imaging data used in this study are not publicly available due to ethical and privacy considerations involving neonatal subjects. To minimize any risk of privacy breaches, the raw MRI data cannot be shared outside the institution. However, de-identified data related to image quality analyses, objective metrics, and aggregated results are available from the corresponding author upon reasonable request, subject to institutional and data-sharing policies.

